# Fusion of magnet resonance imaging and electronic health records promotes the multimodal prediction of postoperative delirium

**DOI:** 10.1101/2025.02.20.25322564

**Authors:** Niklas Giesa, Andrea Dell’Orco, Michael Scheel, Carsten Finke, Felix Balzer, Claudia Doris Spies, Maria Sekutowicz

## Abstract

The role of individual brain morphometry, derived from clinical imaging, for the analysis as well as the early prediction of postoperative delirium (POD), as a severe complication after a surgery, is currently under-explored. We extensively analyzed patient trajectories of magnet resonance imaging (MRI) and electronic health records (EHRs) for two POD definitions covering 557 and 201 patients. Age-adjusted correlations with linear mixed-effect models identified middle temporal and superior temporal cortical thickness as well as thalamus and brain stem volumes. We trained highly non-linear multi-layer perceptrons (MLPs) on EHRs, MRI measures, and the combination of both as multimodal fusions. MLP models achieved high performance metrics, as are under receiver operating characteristics (AUROC) values up to 86%, outperforming baselines. Multimodal fusion was especially beneficial for 645 less critically ill patients. MLP model weights for this cohort focused on cerebral atrophy measures of higher order cortical areas, such as the temporalpole -, superiofrontal gyrus-, and the insula. Our results pave the way for the far unrecognized potential of clinical MRI features for early multimodal predictions of POD.

## 1. Introduction

Delirium is a severe condition with disturbance in major brain functions such as consciousness, cognition, and attention [1–3]. Following a major surgery, postoperative delirium (POD) is associated with adverse outcome such as death and prolonged hospitalization. Prevalence rates span from 5 to 52% [4–7]. Multiple predisposing and precipitating risk factors have been identified [4,6]. The latter include adjustable factors involving the current surgical procedure, while predisposing factors may be preexisting, such as cognitive impairment or advanced age [8,9].

POD is a highly distressing condition with an acute onset and heterogenous levels of vigilance, neuropsychological, and psychotic symptoms [4]. Temporal fluctuations of symptoms in presence and severity are patient-specific and demand a close monitoring and assessment [10]. An individual risk estimation remains challenging due to multifactorial interacting etiologies. These may lead to a common but heterogenous core syndrome that is dependent on brain resilience to acute distress. In the state of delirium, this vulnerability has been associated with cerebral atrophy [11] and is positively correlated with long term cognitive decline [12]. Multiple neuronal mechanisms may be facilitated by preexisting neuroanatomical changes and thus contribute to delirium pathophysiology as a result of neuronal dysfunction and network disintegration [13].

Neuroanatomical pre-morbidity in POD patients has been identified as decreased white matter integrity and increased gray matter atrophy [14–16]. Due to the large range in prevalence rates and in absence of a general pathogenic model, previous studies have been restricted to specific patient cohorts at risk, such as elderly patients undergoing major surgical procedures [14,17]. As a result, the previously identified structural brain changes may be age-specific or limited to patients with preexisting cognitive impairment [15]. A few experimental studies have generated machine learning (ML) prediction models for delirium [18,19] which excel over standard statistics discovering non-linear relationships. However, prediction results seldom translate into clinical practice. Either they are limited to study data over routinely collected records, or prediction models learn from non-generalizable cohorts [20–23].

Outcome prediction across medical fields benefits from the ability of ML to leverage diverse data modalities [24,25]. Mohsen et al. introduce different types of fusion strategies occurring either early in the feature space or later when outputting prediction probabilities [26]. The additional value of these fusion strategies combining electronic health records (EHR) and magnet resonance imaging (MRI) modalities for POD predictions remains underexplored [27,28].

To our knowledge, there are no studies utilizing MRI data out of routine clinical practice to investigate premorbide structural brain changes in POD. Thus, this study aims to proof the applicability of neuroanatomical features extracted from preoperative clinical MRI on POD predictions. Secondly, the combination of such features with EHRs is systematically investigated on general surgical cohorts. Interpretation of results from different multimodal ML techniques was augmented by advanced linear mixed-effect models (MEM). MEMs corrected for covariates, such as age or brain lesions, allowing to draw conclusions about the information value of specific clinical MRI and EHR-features allowing insights into a general pathomechanisms of POD.

## 2. Methods

### 2.1. Ethics Vote and Institutional Review Board

This study was approved by the Ethics Committee of the Charité Universitätsmedizin – Berlin (EA2/024/18) and followed the Declaration of Helsinki. Patient consent for general research purpose was covered by the patient treatment contract. Specific retrospective analysis of patient data approved the IRB EA2/024/18.

### 2.2. Study Population, Endpoint Definitions, and Data Extractions

Postoperative delirium was defined as a dichotomous target variable (1 = delirious, 0 = non-delirious / controls) according to two endpoint definitions with two complimentary cohorts and separate analyses. Primarily, for one endpoint (scoPOD), the CAM-ICU and Nu-DESC [29] were used postoperatively to label patients as delirious (Nu-DESC >0 or positive CAM-ICU) or non-delirious (Nu-DESC= 0 or negative CAM-ICU). Since POD may often be under-recognized, a second endpoint (medPOD) was defined based on agitation and treatment as proxy to diagnosis: Patients who were given antipsychotic medication after surgery and scored positively on the RASS > 1 were classified as delirious. Non-agitated patients (RASS = 0) who had no antipsychotic medication after surgery until discharge were seen as non-delirious.

Error! Reference source not found.a, displays inclusion criteria for the described cohorts. We included all adult patients (aged >= 18), who underwent surgery between 2017 and the end of 2022 at one of the three sites of our medical center, if the estimated surgery duration was >= 1 hour. We extracted sample clinical MRI scans for a random sample of included patients. MRI headers were filtered for head-scan, acquisition time and presence of T1-weighted MPRAGE sequences. In case of multiple preoperative MRI scans during one hospital stay, we selected only one closest to surgery begin. As a result, we investigated 645 scans for 557 patients assigned to endpoint scoPOD and 224 scans for 201 patients regarding the endpoint medPOD.

Error! Reference source not found.b depicts our data extraction and - analysis plan. The picture archiving and communication system (PACS) provided preoperatively scanned cranial MRIs. The clinical information system (CIS) accessed pre- and intraoperatively collected EHRs in addition to two defined postoperative endpoints [30]. EHRs were de-identified and stored in a Data Warehouse (DWH) [31,32]. In the end, we harmonized data from different sources and applied ML techniques.

### 2.3. Data Preprocessing

Due to the nature of routinely collected EHRs potentially following skewed distributions including outliers [33], we aggregated clinical parameters with robust summary statistics as median, 10^th^ – and 90^th^ percentile, alongside the mean. Summarizations were performed separately for the pre- and intraoperative phase. Our initial feature space covered clinical parameters that were available for at least 10% of patients. Laboratory values (lymphocytes, CRP, etc.), medications (propofol, norepinephrine, etc.), vital signs (heart rate, spo2, etc.), and device settings (FiO2, PEEP) were covered resulting in 133 features (see **Extended Table A1/A2** in Supplementary A). All data were normalized via z-transformation with statistics from the training set.

Clinical billing codes were used to encode the presence of diagnosis in form of ICD [34]. We characterized our two analysis cohorts with these codes, but omitted the information for training ML models or endpoint definitions due to uncertain documentation times [35].

### 2.4. Statistical analysis

We analyzed the discriminative power of numerical variables towards the binary endpoint with robust non-parametric Mann-Whitney U (MWU) test statistics [36]. The MWU U statistic directly translates to the area under the curve (AUC) [37]. We report the AUC-0.5 with 0 as a chance-level discrimination, values near 1 as a strong positive effect, and values near -1 as a strong negative effect size. For describing associations of numerical clinical parameters with age, we implemented robust Spearman’s rank correlations [38]. The Spearman correlation coefficient (ρ) defined the effect strength similar to the previous one with levels of 0, near 1 or -1.

Due to the prominent role of age as a confounder in MRI-based brain morphometry measures, we configured multivariate linear mixed-effect models (MEMs) [39,40] that correct for this relationship. We configured MEMs with varying intercepts for single feature effects as *C*(*POD*) ∼ *age* + *feature* + 1|*patient*. Here, the POD endpoint functions as the dependent variable, age is integrated as fixed effects in conjunction with the feature of interest. Due to patients potentially having multiple surgeries, we integrated the patient identifier as a random effect. We report an age-adjusted p-value and a corresponding coefficient β(f) indicating feature effect directions.

Statistical significance was assessed using false discover rate (FDR) corrections (alpha = 0.05) reducing the risk of type I errors falsely rejecting the null-hypothesis [41]. For reporting the effect strengths of binary variables with POD, we used the odds ratio (OR) on a logarithmic scale as ln(OR) with large deviations from 1 indicating strong effects [42].

### 2.5. MRI analysis

All images were first converted from DICOM to NIfTI format using dcm2niix and then segmented using the FreeSurfer v7.4.1 recon-all pipeline [43] Morphometry measures such as thicknesses, volume, area, and curvature for each region of interest (ROIs) were computed from the brain segmentations and the cortical parcellation using the Desikan-Killiany atlas [44]. We performed a visual quality control of all segmentation results to control for possible missegmentation due to both the administration of gadolinium-based contrast agents or to anatomical aberrations, such as general atrophy, tissue lesions including tumors. In the case of any brain abnormality restricted to one hemisphere the healthy hemisphere of each subject’s brain was chosen to be included in the analysis. For healthy brains, one hemisphere was selected randomly for analyses. In the scoPOD (n=645) cohort, 358 right – and 287 left hemispheres were selected. For medPOD (n=224), 91 right – and 133 left sides were chosen. To account for the effect of intracranial volume in the volume estimate, the volumes were normalized by dividing them by the estimated total intracranial volume (eTIV). The final MRI-related features set was composed of 184 volumes either from FreeSurfer segmentation or parcellation, 70 thickness features, and 72 area features.

### 2.6. Machine Learning and Fusion Strategies

We trained three different ML techniques comprising logistic regression (LR), gradient boosted trees (BT), and multi-layer perceptron (MLP) architectures [45] that were configured for our POD prediction task. While LR assumes linear relationships between the logarithmic odds of predictors and the outcome, BT and MLP can be fitted to highly non-linear problems. BT, as a type of ensemble model, constructs inter-connected decision trees (DT) that partition the feature space sequentially (branches) according to self-leaned rules [46]. MLP models function as universal approximators [47] stacking so-called perceptrons (nodes) on layers while connecting each node on one layer with nodes from the subsequent one, so forming a network.

For handling MRI as well as EHR modalities, we deployed two fusion strategies [48]. In “early fusion”, we enhanced our input feature space by simply ingesting selected measures from both types. For the “late fusion” strategy, we trained two models in parallel, each for one modality (MRI or EHR), and combined the prediction outputs in the end (see **Extended Figure B1** in Supplementary B). For BT and LR, model outputs (probabilities between 0 and 1) were simply mean-averaged. In the late fusion MLP, a linear layer combines predictions from both models to a single one while learning model weights for both networks via backpropagation, also known as “joint-fusion” [24,48]. As a reference, we also trained completely separate models for MRI and EHR features only.

### 2.7. Model Configuration, Training, and Validation

For finding the best model configurations (hyperparameters), we implemented a 3x3 nested cross-validation (CV) [49] approach where all data is split three times in three equally-sized partitions (folds). While two folds (66% of data) are used for training, models are iteratively evaluated on one hold-out fold (33% of data). Different sets of parameters are exhaustively validated via a Grid-Search [50] process on the inner-nested CV process and then applied to the outer-nested one. Besides, 1000x bootstrapping, as sampling with replacement on the outer validation fold, enabled us to estimate the 95% confidence interval (CI) [51] of validation results.

The area under (AU-) the receiving operating characteristics (-ROC), and the precision recall curve (-PRC) were used to evaluate performances. [37]. As a cost function, we configured a binary cross-entropy (BCE) loss that models aimed to minimize [52]. The final parameter space included regularization techniques, like BT pruning or L1-norm penalty for MLP and LR, in addition to general configurations (see **Extended Table B2** in Supplementary B). We trained models with subsets of features for different adjusted p-values thresholds (see **Extended Table A3** in Supplementary A).

## 3. Results

### 3.1. Cohort Characteristics

**Table 1** displays cohort characteristics for scoPOD and medPOD based on extracted EHR variables, the POD prevalence was 18.44% and 21.43%, respectively. We cite p-values as discriminating against POD cases and controls per endpoint. Patients who suffered from POD were older (65.01±14.04 years as mean±sd for scoPOD, 62.08±13.49 years for medPOD) than controls (58.02±16.43 years for POD, 56.85±16.02 years for medPOD, p-values = 4.94E-04 and 1.45E-03). The medPOD cohort covered slightly younger patients than the first one scoPOD (Δ of mean age = 1.92 years), both included more male patients than females (78 and 69%).

**Table 1:** Descriptive cohort characteristics for two POD endpoints. Descriptive statistics are displayed as mean±sd [1st,2nd,3rd] quartile for numerical variables. For binary variables, the fraction of positive samples from all (n) are cited followed by the odds as (pos/neg samples). Adjusted p-values are derived from linear mixed-effect models (MEMs) incorporating age and the variable of interest as fixed effects, patient groups as random effects and POD as the independent variable. We highlight significant results with boldness and asterisk* according to a FDR corrected alpha level.

|  | Endpoint 1: scoPOD (n=645) |  | Endpoint 2: medPOD (n=224) |  |
| --- | --- | --- | --- | --- |
|  | Descriptive Statistics [all] | Adjusted P-Value [POD] | Descriptive Statistics [all] | Adjusted P-Value [POD] |
| **General Information** |  |  |  |  |
| Age (years) | 59.32±16.23<br>[50,62,72] | **4.94E-04\*** | 58.03±15.61<br>[48,60,70] | 1.45E-03 |
| Sex (male/female) | 0.78 (510/135) | 1.90E-01 | 0.69 (155/69) | 2.00E-01 |
| Number of Surgeries | 2.27±3.32<br>[1,1,2] | 1.00E-01 | 1.94±2.23<br>[1,1,2] | 2.10E-01 |
| Length of Hospital Stay (days) | 31.45±3.10<br>[10.08,17.08,35.31] | 1.29E-01 | 34.25±4.80<br>[12.12,20.92,36.13] | 1.22E-01 |
| Length of Anesthesia (hours) | 3.10±3.07<br>[1.67,2.49,3.59] | 2.62E-02 | 5.14±2.97<br>[2.22,3.19,4.43] | 1.28E-02 |
| Length of Surgery (hours) | 1.52±1.16<br>[0.70,1.25,1.99] | 2.93E-01 | 1.98±1.34<br>[0.96,1.60,2.67] | 1.64E-01 |
| Length of Recovery Room Stay (hours) | 4.82±2.66<br>[1.21,1.89,2.99] | **3.31E-05\*** | 5.14±2.78<br>[1.18,1.91,3.23] | 4.42E-01 |
| **Clinical Assessment** |  |  |  |  |
| Urgency Class N | 4.24±1.79<br>[4,5,5] | 1.94E-01 | 4.12±1.44<br>[4,5,5] | 1.11E-02 |
| ASA Status | 1.93±1.22<br>[1,2,3] | **4.61E-04\*** | 2.06±1.34<br>[1,2,3] | **9.14E-05\*** |
| SOFA | 4.25±3.18<br>[1.83, 3.70, 6.51] | 1.86E-03 | 4.95±3.22<br>[2.73, 4.24, 7.18] | 4.52E-04 |
| RASS | -0.66±1.00<br>[-0.97, -0.36, 0.00] | 7.29E-02 | -0.90±1.14<br>[-1.37, -0.60, -0.09] | **2.97E-04\*** |
| **Type of Surgery** |  |  |  |  |
| Neurosurgical | 0.56 (363/282) | 5.44E-02 | 0.60 (134/90) | **9.79E-08\*** |
| Visceral | 0.18 (119/526) | 4.69E-01 | 0.21 (48/176) | 3.94E-01 |
| Anesthesia | 0.07 (46/599) | 1.07E-02 | 0.06 (13/211) | 4.41E-01 |
| Urology | 0.02 (15/630) | 2.68E-03 | 0.02 (4/220) | 1.24E-01 |
| Cardiac | 0.01 (9/636) | 7.47E-02 | 0.04 (8/216) | 3.89E-01 |
| Orthopedic | 0.05 (32/613) | 3.34E-01 | 0.07 (14/210) | 6.76E-02 |
| Otorhinolaryngology | 0.05 (33/612) | 2.58E-01 | 0.02 (4/224) | 2.58E-03 |
| **Predisposing Risk** |  |  |  |  |
| Cardiovascular | 0.56 (363/282) | 1.89E-02 | 0.57 (128/96) | 1.06E-02 |
| Non-Smoking | 0.31 (197/448) | 7.51E-01 | 0.40 (89/135) | 1.05E-01 |
| Smoking | 0.04 (24/624) | 4.54E-01 | 0.05 (12/212) | 2.42E-01 |
| Other Comorbidities | 0.02 (10/635) | 6.70E-02 | 0.02 (4/220) | 1.76E-01 |
| Drinking | 0.01 (5/640) | 2.38E-01 | 0.01 (2/222) | 6.08E-03 |
| **MRI Screening** |  |  |  |  |
| Tumor | 0.18 (114/531) | 7.71E-01 | 0.20 (45/179) | 6.88E-02 |
| Atrophy | 0.07 (42/603) | 9.48E-01 | 0.08 (18/206) | 3.25E-01 |
| Lesion | 0.43 (279/366) | 9.65E-01 | 0.53 (119/105) | 6.10E-01 |
| Contrast Agent | 0.79 (512/133) | 8.86E-01 | 0.82 (184/40) | 7.24E-01 |
| **ICD Diagnose** |  |  |  |  |
| Any Malignancy | 0.50 (321/324) | 2.68E-01 | 0.51 (105/109) | 1.99E-01 |
| Renal Disease | 0.37 (241/524) | **3.19E-07\*** | 0.43 (97/127) | **3.24E-06\*** |
| Metazoic Tumor | 0.28 (179/466) | 9.88E-01 | 0.29 (66/158) | 4.76E-02 |
| Delirium | 0.19 (118/517) | **3.61E-26\*** | 0.30 (68/156) | **2.34E-28\*** |
| Hemiplegia Paraplegia | 0.20 (129/516) | 3.63E-01 | 0.28 (62/162) | 4.55E-01 |
| SIRS | 0.14 (92/543) | **7.96E-05\*** | 0.21 (46/178) | **2.99E-20\*** |
| Sepsis | 0.12 (79/556) | 1.10E-03 | 0.17 (38/186) | **5.94E-13\*** |
| Dementia | 0.02 (13/632) | 4.92E-03 | 0.04 (8/216) | 8.13E-01 |
| Liver Disease | 0.01 (6/639) | 2.72E-03 | 0.02 (4/220) | 5.20E-03 |
| Schizophrenia | 0.02 (10/635) | 3.87E-03 | 0.02 (4/220) | 1.33E-01 |

**Table 2:**
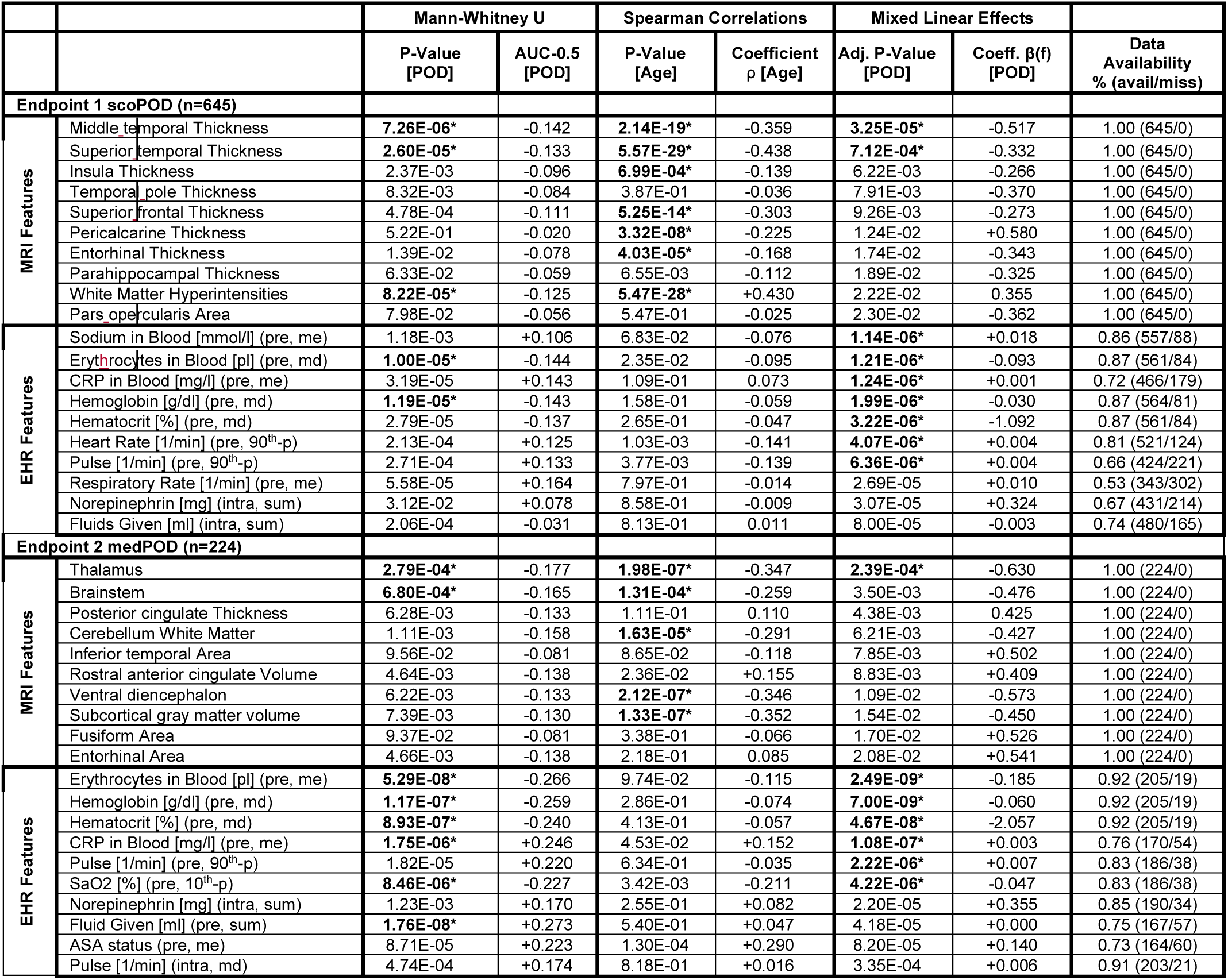
Single feature importance for MRI and EHR features per endpoint. Univariate results from Mann-Whitney U (MWU) test statistic define discriminability of [POD] endpoints with unadjusted p-value and AUC-0.5 as effect size. Spearman Ranks provide correlation coefficient (ρ) calculated between feature values and [Age] including p-value under the null- hypothesis of zero-coefficients. Linear mixed-effect models (MEM) were fitted with [POD] as dependent variable, feature values as fixed effects, patient-mri hierarchy as random effects. MEMs provide age-adjusted p-values and a corresponding coefficient β(f). Data availability shows the fraction of available feature values from all, adjacent to the odds of (available/missing) values. EHRs must be aggregated with either the sum, median (md), mean (me) for preoperative (pre) or intraoperative (intra) time phases.

We found significant differences for the length of recovery room stay for scoPOD (5.18±2.42 hours for POD, 2.81±3.28 hours for controls, adjusted p-value = 3.31E-05). Patients included in medPOD stayed 0.31 hours longer in the recovery room than scoPOD patients. While patients had decreased physical status in the medPOD cohort (Δ of mean ASA = 0.13), they were more prone to sequential organ failure (Δ of mean SOFA = 0.70) and less agitation (Δ mean RASS = 0.24) compared to scoPOD. For medPOD, we found statistically significant differences for patients’ physical status (ASA 2.12±1.21 POD, 1.82±1.69 controls, p = 9.14E-05) and RASS (-1.08±1.16 POD, - 0.81±1.12 controls, p = 2.97E-04). The RASS was present preoperatively for 12% and 35% for scoPOD and medPOD respectively indicating the fraction of patients that were admitted to ICUs before a surgery.

In both cohorts, the most prominent surgical procedure was neurosurgery, reaching a significant difference for POD labels in medPOD (ln(OR)= 1.71, p = 9.79E-08). There was no strong evidence that manual MRI screening properties, such as visual atrophy, were discriminative for POD (see **Table 1**). The presence of ICD encoded renal disease (ln(OR) = 1.13, p-value = 3.19E-07 for scoPOD, ln(OR) = 1.47, p-value = 3.24E-06 for medPOD) and SIRS (ln(OR) = 0.92, p-value = 7.96E-05 for scoPOD, ln(OR) = 2.36, p-value = 2.99E-20 for medPOD) were significantly more often found in patients with POD for both endpoints. The presence of sepsis was significant for medPOD with ln(OR) = 2.29 and p-value = 5.94E-13. As a sanity check for our two POD definitions, we observed very strong correlation with ICD encoded delirium (ln(OR) = 2.10, p-value = 3.61E-26 scoPOD, ln(OR) = 2.92, p-value = 2.34E-28 medPOD). More information about surgical procedures is included in the appendix (**Extended Table B3** in Supplementary B).

### 3.2. MRI and EHR Single Feature Importance

Analyzing importance of MRI-derived morphometry features, the middle temporal- and superior temporal thickness expressed significant strong univariate correlation with POD (MWU p-value =7.26E-06, AUC-0.5 = -0.142; p-value =2.60E-05, AUC-0.5 = - 0.133) as well as with age (Spearman p-value = 2.14E-19, ρ = -0.359; p-value = 5.57E- 29, ρ = -0.438) regarding scoPOD (see **Table 1**, **Figure 2**a). Decreased levels of cortical thickness resulted in increased probabilities of POD (negative MWU effect size as AUC-0.5) and occurred further in elderly -, then in younger patients (negative Spearman effect size as ρ).

**Figure 1:**
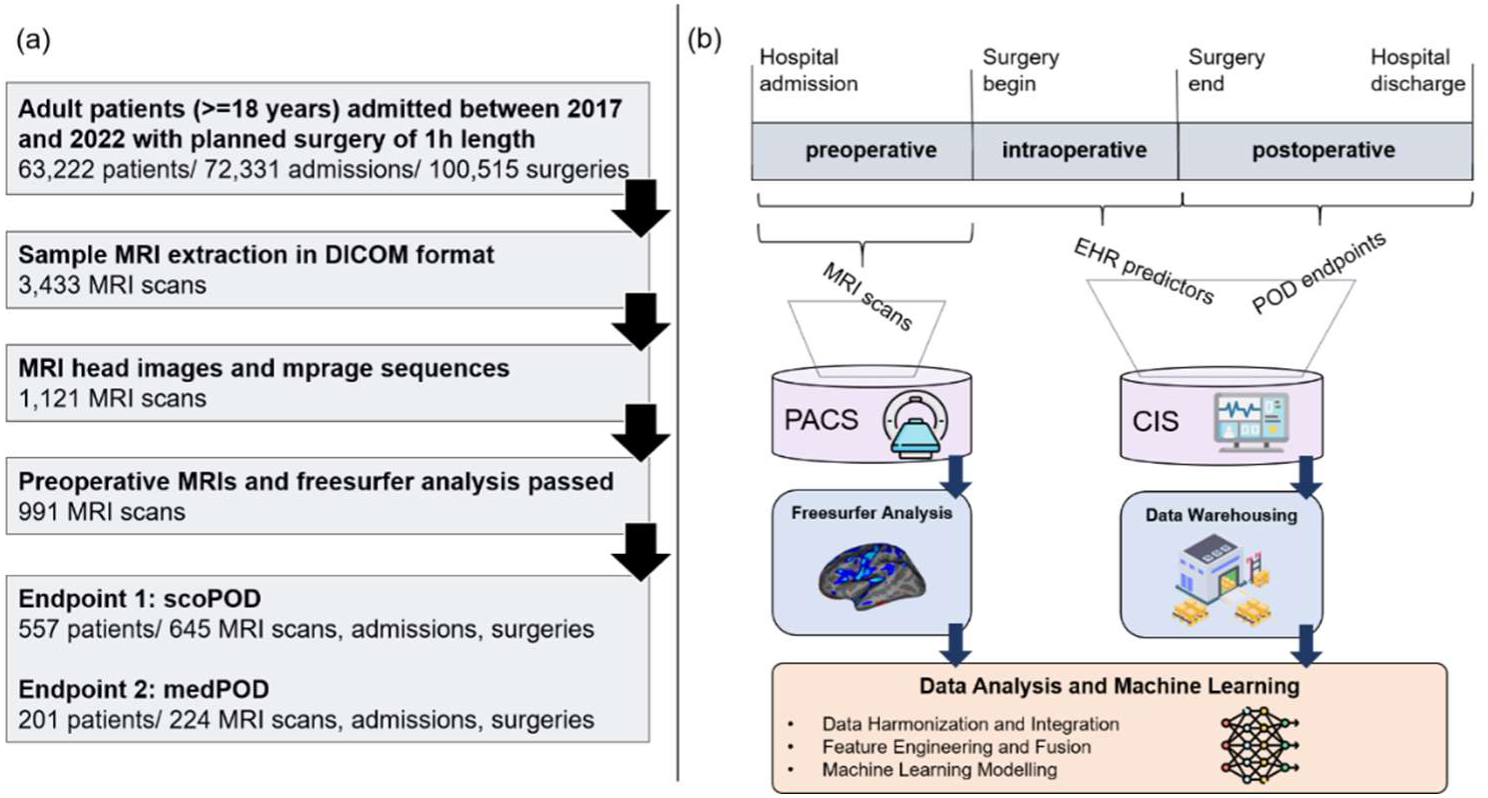
Inclusion criteria for endpoint definitions (a), methodology and data flow (b). We included data of adult patients between 2017 and 2022 with estimated surgery length of at least one hour. Posthoc MRI headers were filtered for head-scan and MPRAGE sequences and scanning time. a) Preoperative MP-RAGE sequences that did not pass automated segmentation (e.g. extreme movement) were excluded (shown in a). We divided the hospital stay into pre-, intra-, and postoperative time phases. MRI scans were extracted from the picture archiving and communication system (PACS), EHR data were extracted from the clinical information system (CIS) for integrated data analysis and the application of machine learning (shown in b).

**Figure 2:**
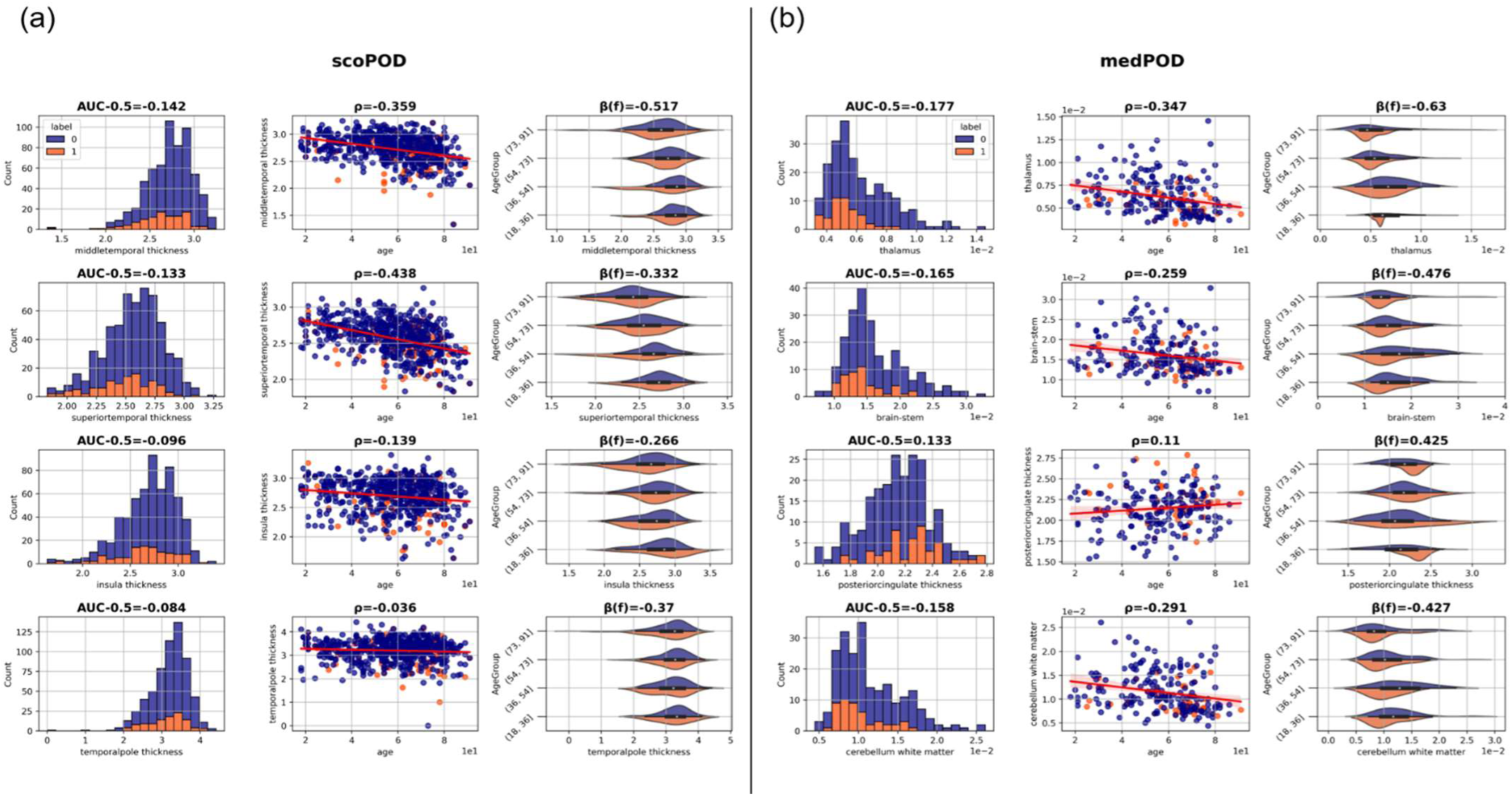
Top 4 single MRI feature importance for endpoint scoPOD (a) and medPOD (b). We display most significant results according to an age-adjusted p-value derived from mixed- linear-effects models (MLEMs). For each panel (a) or (b), the first column displays the distribution of feature values for POD cases (label=1, orange) and controls (label=0, blue) including AUC- 0.5 as an effect size derived from Mann-Whitney U (MWU) statistics. The second column shows the correlation between the feature and age providing the Spearman Rank correlation coefficient ρ. The last column depicts age-group specific differences in distributions of POD cases and controls for each feature citing the corresponding coefficient β(f) retrieved from MLEMs.

Multivariate MEM analysis showed that cortical thickness parameters for the middle as well as the superior temporal cortex remained significantly associated with scoPOD when adjusting for age (adj. p-value = 3.25E-05, β(f) = -0.517; adj. p-value = 7.12E-04, β(f) = -0.332). The trend of decreased cortical thickness in POD was preserved, even when dividing patients into equally-sized age groups (negative MEM coefficient β(f), see **Figure 2**a). Apart from thickness, the measure of white matter hypointensities expressed significant univariate effects on scoPOD (MWU p-value =8.22E-05, AUC-0.5 = -0.125) and age (Spearman p-value = 5.47E-28, ρ = 0.430).

MWU analysis of EHRs highlighted preoperative measures of anemia (hemoglobin: p-value = 1.19E-05) and infection parameters (erythrocytes: p-value = 1.00E-05). These and additional ones were significant after age-corrections (MEM erythrocytes: adj. p-value = 1.21E-06; hemoglobin: adj. p-value = 7.00E-09, hematocrit adj. p-value = 3.22E-06; CRP: adjusted p-value = 1.24E-06). Age-corrected MEMs also detected significant measures of centrally - and peripherally measured heart rate (heart rate: adj. p-value= 4.07E-06; pulse: adj. p-value= 6.36E-06) for this endpoint.

For medPOD, subcortical MRI features, such as thalamus and brainstem volume were significantly associated without age correction (thalamus volume, p-value =2.79E-04, AUC-0.5 = -0.177; MWU; brainstem volume p-value =6.80E-04, AUC-0.5 = -0.165; MWU). After correcting for age, the thalamus volume remained significant (adj. p-value = 2.39E-04) (see **Table 1**, **Figure 2**b). Volumetric measures of the subcortical gray volume expressed high correlations with age (Spearman p-value = 1.33E-07, ρ = -0.352). MWU analysis detected more EHR correlations for medPOD than for scoPOD. MEM for medPOD also revealed blood parameters like lowered levels of erythrocytes (adjusted p-value = 2.49E-09, β(f) = -0.185), hemoglobin (adjusted p-value = 7.00E-09, β(f) = - 0.060), hematocrit (adjusted p-value = 4.67E-08, β(f) = -2.057), and increased CRP (adjusted p-value = 1.08E-07, β(f) = 0.003) (see **Table 1**).

### 3.3. Machine Learning Results

We assessed the quality of trained LR, BT, and MLP models with AUROC and AUPRC metrics for both endpoints. Best results for scoPOD were achieved by a late fusion MLP (AUROC 0.735 [0.726, 0.744] as mean, [95% CI]; AUPRC 0.456 [0.411, 0.472]), outperforming LR (AUROC 0.705 [0.695, 0.715]; AUPRC 0.404 [0.389, 0.420]) and BT (AUROC 0.722 [0.712, 0.32]; AUPRC 0.450 [0.436, 0.468]) within the same fusion type (see **Figure 3**a).

**Figure 3:**
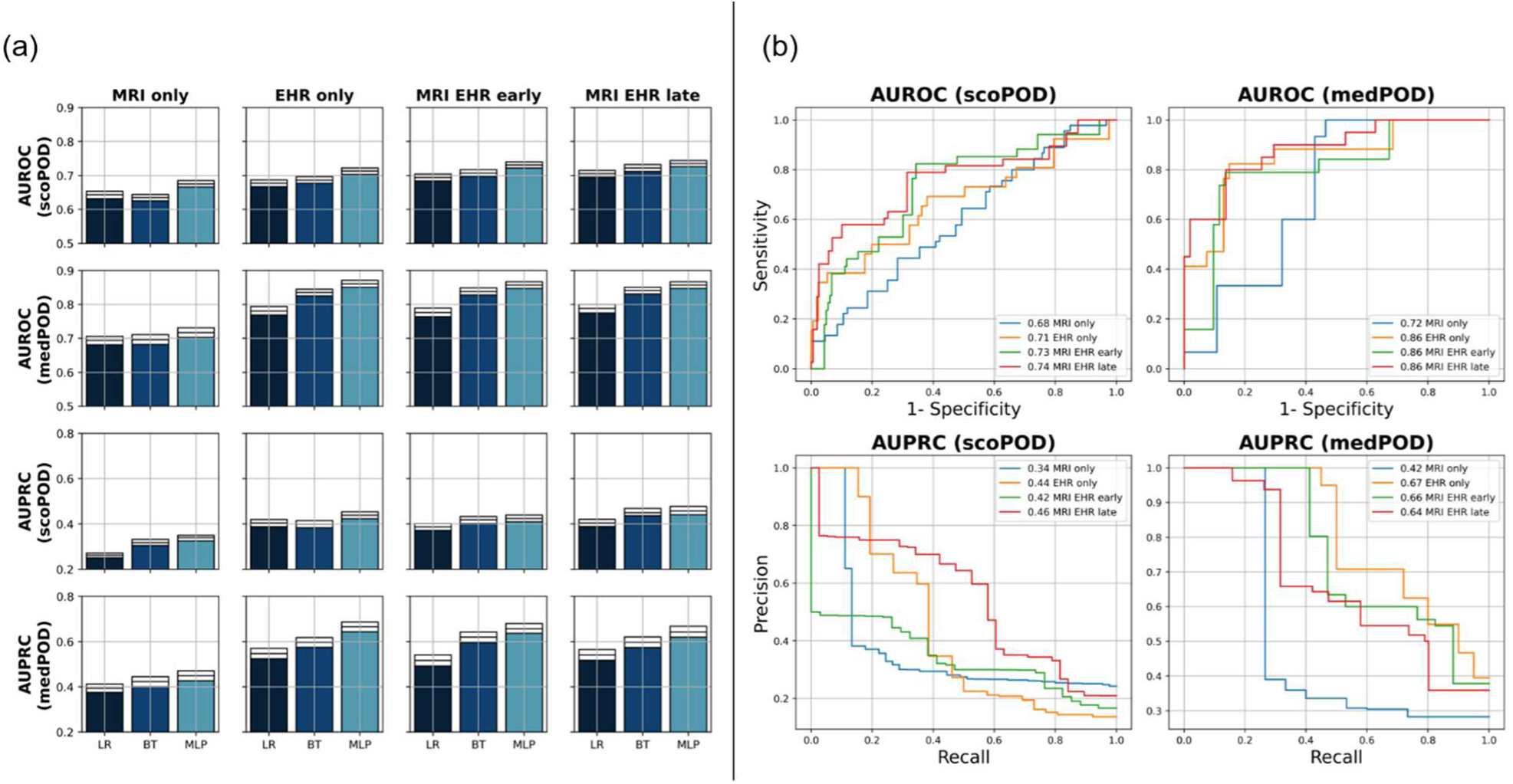
Performance metrics across machine learning models and fusion types (a) or for best performing model (b). Area under (AU-) the receiver operating characteristics (- ROC) or the precision recall curve (-PRC) for trained and 3x cross-validated machine learning methods applied to predict endpoint scoPOD (n=645) or medPOD (n=224). Gradient boosted trees (BT), logistic regression (LR), and multi-layer perceptrons (MLPs) are included. Panel a shows metrics across models and fusion types either including magnetic resonance imaging features (MRIs), or electronic health records (EHRs) only, or combining these two modalities (early, late). 95% confidence intervals are drawsn as white boxes in (a) on top of bars calculated on 1000x bootstrapped validation folds. In panel b, trained MLPs are selected to draw AU curves showing mean performances across validation sets.

MLPs were superior to LR and BT for all fusion types (see **Extended Table B4 in** Supplementary B). We observed overlapping CIs of AUROC between late- and early fusion of MLPs for scoPOD ([0.726, 0.744] vs. [0.722, 0.740]), but distinct differences to MLPs trained with one modality only (EHR only [0.703, 0.721], MRI only [0.666, 0.685]). Combined fusion models benefited especially from the inclusion of EHRs.

Performances of models predicting medPOD were elevated in contrast to results for scoPOD (see **Figure 3**a). Best performance was achieved by the MLP ingesting EHR features only (AUROC 0.861 [0.851, 0.871]; AUPRC 0.665 [0.644, 0.687]). Here, the confidence was decreased due to similar metrics ranges yielded by combined fusion MLPs like early - (AUROC [0.847, 0.860]; AUPRC [0.636, 0.679]), or late fusion (AUROC [0.847, 0.867]; AUPRC [0.619, 0.668]).

MLPs also showed elevated validation metrics for medPOD in contrast to other ML methods. The inclusion of EHRs was exquisitely important for successful prediction of medPOD cases.

In **Figure 3**b, corresponding AUROC and AUPRC curves describe model behaviors under varying prediction thresholds. Curves confirm that late fusion was favorable for the first endpoint scoPOD with a sensitivity of 0.81 and a specificity of 0.63 at the threshold where their sums maximize. MLPs trained solely on EHRs could even exceed these metrics with 0.81 sensitivity and 0.82 specificity predicting the medPOD endpoint.

### 3.4. Model Interpretation

We read model weights (MW) from our best performing MLPs per endpoint to interpret the post-hoc feature importance. The best late fusion scoPOD MLP focused its weights inside the EHR model on intraoperative measurements like tidal volume (abs MW = 0.288) and preoperative measures of hydration such as albumin blood levels (abs MW = 0.283) and erythrocytes count (abs MW = 0.2). Levels of infectious parameter C-reactive protein (CRP) (abs MW = 0.132) the amount of given norepinephrine (abs MW 0.192), or the physical status (ASA abs MW = 0.169) were also predictive.

Highest model weights, derived from the late fusion MLP ingesting MRI features for scoPOD, were assigned to temporal pole thickness (MW=0.565), superior frontal gyrus (MW=0.523), and insula thickness (MW=0.504). These features also showed univariate feature importance examined by previous MWU statistics with effect strengths of 0.084 (p-value=8.32E-03), -0.111 (p-value=4.78E-04), and -0.096 (p=2.37E-03) AUC-0.5 (see **Extended Table B5** Supplementary B, **Figure 4**). The middle temporal – and superior temporal gyrus showed high MWU importance but where ranked relatively lower by our trained MLP.

**Figure 4:**
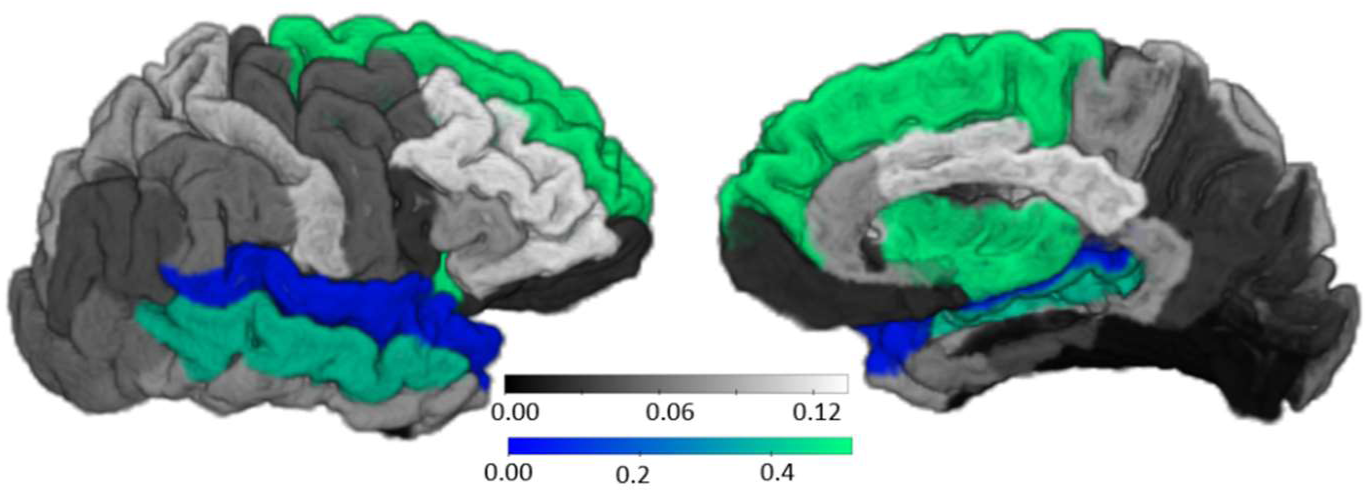
Model importance of MRI-features for scoPOD. The absolute value of AUC-0.5 derived from MWU (grey scale) and absolute model weights (MW) from best MLP classifier (blue-green scale) trained with MRI only are shown. Anatomical segmentation across right hemisphere according to DKT-Atlas is displayed from temporal and medial. Overlay colored green-blue indicates model weights. Underlaying absolute AUC metrics for blended areas are: superior frontal thickness 0.111, insula thickness 0.096, middle temporal thickness 0.142, superior temporal thickness 0.133, temporal pole thickness 0.084.

The best MLP classifying medPOD relied purely on EHRs. Highly weighted features were the mean blood pressure (abs MW = 0.186), or the volume of given fluids (abs MW = 0.183). The MLP focused on intraoperative measures such as the application of norepinephrine (abs MW = 0.172), the tidal volume (abs MW = 0.166), and heart frequency (abs MW 0.142). Preoperative CRP levels (abs MW = 0.158), preoperative physical status (ASA abs MW = 0.120), or hydration measures such as hematocrit (abs MW 0.135) and erythrocyte count (abs MW 0.133) had predictive values as well (see **Extended Table A4** in Supplementary A). Additional MRI weights for medPOD provided by the unimodal MLP are covered in the supplement (see **Extended Table B5** in Supplementary B). We assessed potential model prediction biases by analyzing the relationship between key covariates and model raw output probabilities for each surgery, confirming the absence of these biases (see **Extended Figure B6** in Supplementary B).

## 4. Discussion

This study proves that POD predictions benefit from the fusion of clinical preoperative MRI based morphometry with EHRs. By iteration of data fusion strategies and ML models we are the first to demonstrate that neuroanatomical features from routinely collected MRIs promote predictive models to capture both localized brain vulnerabilities and multimodal risk factors that contribute to POD. Cortical thickness parameters of the temporal gyrus as well as subcortical volumetric measures, including the thalamus and brainstem volume contributed to successful POD-prediction.

In a general surgical population, we defined two POD endpoints, which were highly correlated with ICD encoded delirium, but reflect distinct subgroups with key differences in clinical and surgical characteristic. POD definitions based on standard assessment tools (scoPOD) covered a wider range of surgical interventions while medPOD patients showed a significant association between neurosurgery and POD. The medPOD definition included more critically ill patients as evidenced by higher prevalence of ICU stays before surgery, higher degree of critical illness, and systemic inflammation.

Findings of scoPOD associated cortical thickness parameters in the temporal lobe, involved in memory, attention, and higher-order executive functions, align with literature linking temporal cortex atrophy to delirium [8]. These cortical features outperformed EHR predictors, suggesting that brain-specific vulnerability, reflected by morphometry of the temporal gyrus, is primary driving POD in less critically ill populations. When critical illness is more predominant and patients are in need of antipsychotic pharmacotherapy, subcortical MRI measures outperform structural measures of the temporal gyrus, emphasizing the importance of fronto-striato-thalamic circuits in disorders of consciousness and the emergence of psychotic symptoms [53]. However, POD predictions with EHR features only outweighed the influence of structural brain features resulting from the clinical complexity of critical illness. Here, preoperative anemia, hydration and infection proxies, that are significantly associated with POD in univariate tests, showed high value for prediction. In the more general population of our surgical patients, existence of predisposing risk factors such as preoperative infection as well as dehydration may be crucial for correct POD prediction.

In general, the performances of MLP prediction models were evaluated with very high scores up to 86% AUROC and 66% AUPRC applying fusion strategies. Huang et al. found that related work mostly used early fusion for multimodal integration of data while imaging data is oftentimes processed by pixel-wise convolutional networks [48]. Guidelines suggest empirical investigation of variations to which we adhere by including stand-alone model variants as well as early- and late fusion [26,48]. Xue et al. investigated AKI-related hippocampal damage derived from clinical imaging by trained and interpreted POD prediction models for cardiovascular surgeries. [54]. Instead of keeping the clinical scope narrow, we have provided models that are more generalizable. While previous work successfully trained POD prediction models on EHRs [21,23,27,28], we could show that the combination of data modalities improved performances, especially for a representative less critical ill patients populations with a broader range of surgeries. We used predisposing factors like age, comorbidities, preexisting cognitive impairment, type of surgery and precipatory factors such as lengths of surgery to successfully train our predictive models [27][23]. We encoded these features with robust summary statistics like percentiles to represent temporal fluctuating properties of clinical parameters [10].

To correct for confounders, we applied MEMs focusing on age as a strong covariate [11] increasing statistical power by reducing variance. Overall, MRI features showed stronger age-dependency than EHRs according to Spearman correlations. While age-adjusted MEMs also depicted more significant EHR measurements with respect to both endpoints than MRI features. However, due to highly inter-correlated nature of our data latent noise may persist. For explainable artificial intelligence (XAI) purposes, we preferred directly reading model weights over methods like Shapely or LIME due to their susceptibility to unfavorable effects such as suppressor variables [55,56]. Our multimodal MLP model predicting scoPOD selected most predictive features via an iterative selection approach. Thus MWs, as technical model properties are not suited for drawing clinical implications. Therefore, we provide extensive univariate analyses alongside with model weights to augment clinical interpretations, e.g. age-correction. To further enhance generalizability our prediction results, we conducted analyses for model biases with no significant correlation of age, urgency class, sex, or performed neurosurgery with our model probabilities, suggesting a broad applicability of our prediction results.

Since brain morphometry measures were extracted from clinical MRI assessments manually, we performed quality control to identify alternation from healthy brain anatomy and segmentation inaccuracies. MRI segmentation relied on Freesurfer, originally optimized for healthy brains. Automated morphometry tools, such as Freesurfer, are usually used with non-contrast enhanced (CE) images, however excellent reliability and agreement is reported for the segmentation of T1wCE images [43]. Only one healthy hemisphere was included for patients with lesions, potentially excluding valuable data. In the future, more automated and data-driven segmentation approaches, such as deep learning methods, should be developed to handle scans with potential lesions enhancing prediction robustness. Finally, the sample size, particularly for medPOD, restricts generalizability. Future research should validate these findings in larger, prospective cohorts incorporating external validations.

### 4.1. Conclusion

This study presents the benefits of multimodal fusion models integrating clinical MRI and EHR data leveraging the potentials of modern machine learning in clinical decision making.

## 5. Data-, Code Availability and Reporting

Python code can be accessed via https://github.com/ngiesa/fusion_pod while the individual patient data cannot be shared due to German privacy regulations. We provide comprehensive summary statistics in the Supplement A. We report results according to the “Transparent reporting of a multivariable prediction model for individual prognosis or diagnosis” (TRIPOD) guidelines (see **Extended Table A4** in Supplement A).

## Data Availability

All data produced in the present study are available upon reasonable request to the authors

https://github.com/ngiesa/fusion_pod

## 6. Acknowledgements

The authors acknowledge the Scientific Computing of the IT Division at the Charité -Universitätsmedizin Berlin and of the Berlin Institute of Health Center of Digital Health for providing computational resources that have contributed to the research results reported in this paper.

